# Quantitative Susceptibility Mapping Radiomics with Label Noise Compensation for Predicting Deep Brain Stimulation Outcomes in Parkinson’s Disease

**DOI:** 10.1101/2024.12.26.24319663

**Authors:** Alexandra G. Roberts, Jinwei Zhang, Ceren Tozlu, Dominick Romano, Sema Akkus, Heejong Kim, Mert R. Sabuncu, Pascal Spincemaille, Jianqi Li, Yi Wang, Xi Wu, Brian H. Kopell

## Abstract

Parkinson’s disease patients with motor complications are often considered for deep brain stimulation (DBS) surgery. DBS candidate selection involves an assessment known as the levodopa challenge test (LCT). The LCT aims to predict DBS outcomes by measuring symptom improvement accompanying changes in levodopa dosage. While used in the patient selection process, inconsistent LCT predictions have been widely documented, verified here with Pearson’s correlation *r* = 0.12. Estimating symptom improvement to separate DBS responders and non-responders remains an unmet need. Quantitative susceptibility mapping (QSM) is routinely acquired for pre-surgical planning and depicts the iron distribution in substantia nigra and subthalamic nuclei. Iron deposition in these nuclei has correlated with disease progression and motor symptom severity. A novel QSM radiomic approach is presented using a regression model and features extracted from the pre-surgical targeting acquisition. Noise compensation in training labels improves outcome prediction in regression and classification models. The model predicts improvement in the unified Parkinson’s disease rating scale (UPDRS-III) (*r* = 0.75). Predictive feature maps in deep gray nuclei offer contrast between responders and non-responders. The QSM radiomic approach has potential to improve DBS candidate selection by accurately estimating symptom improvement, eliminating difficult medication manipulation, and avoiding time-consuming evaluations to reduce patient and clinician burden.

## 1. Introduction

Deep brain stimulation (DBS) is a surgery to recalibrate neural circuitry in movement and psychiatric disorders including essential tremor, dystonia, and obsessive-compulsive disorder^1^. DBS in the subthalamic nucleus (STN) is a widespread treatment for advanced Parkinson’s disease (PD) motor symptoms^2^. Selection of candidates for DBS involves assessing responsiveness to levodopa through the levodopa challenge test (LCT). The LCT has shown limitations in estimating improvements, yielding results with large uncertainty^3,4^ and is burdensome to patients and clinicians as it requires multiple assessments and medication withdrawal. Developing a consistent and convenient method for predicting DBS outcomes in PD is an important unmet clinical need. The purpose of this work is to demonstrate the potential of presurgical quantitative susceptibility maps (QSM) to improve patient selection.

From the phase image of multi-echo gradient echo sequences (mGRE), reconstructed QSMs^5^ provide superior contrast in deep gray nuclei during pre-surgical planning^6–8^. QSM is also useful in monitoring disease progression^9^. Recently, deep gray nuclei QSM radiomic features have been shown to improve PD diagnosis ^10,11^, and correlation with both PD motor symptoms^12,13^ and DBS binary outcomes^14^.

Radiomic features represent quantitative information in an image beyond what is discernable to the eye^15^. Such features are generally defined as first-order histogram-based features describing voxel values, second-order features representing joint probability distributions of voxel values, and region of interest (ROI) shape features^16^. Features act as biomarkers across imaging modalities and encode phenotype differences in a variety of pathologies.

The goal of the QSM radiomics model is to preoperatively predict post-surgical UPDRS-III improvement, referred to as “ground truth” or “label” data. As the UPDRS-III metric is measured on an interval scale, a linear regression with the least absolute shrinkage operator (Lasso) penalization technique is proposed using QSM radiomic features to predict DBS outcomes. Specifically, the outcome of interest is the ratio of postoperative to preoperative UPDRS-III scores, known as the UPDRS-III improvement. The UPDRS-III rating contains variability despite good agreement, described in supervised learning as “noise” ubiquitous in human measurements^17^. “Label noise” degrades regression models and supervised learning generally and label noise compensation has been shown to improve model performance^18^. Accordingly, a QSM radiomics Lasso model with noise compensation for DBS outcome prediction is presented.

## 2. Methods

### 2.1 Patient Cohort

This retrospective study was approved by the institutional ethics committee. Informed consent was obtained for forty patients undergoing bilateral STN-DBS. Patients were selected by the department of neurosurgery based on 1) primary PD with good response to levodopa; 2) decreased drug efficacy or motor complications; 3) adverse drug reactions; 4) tremors uncontrolled by drugs. Exclusion criteria were: 1) psychiatric disorders; 2) dementia; 3) diffuse cerebral ischemic lesions on MRI; 4) comorbidities interfering with surgery. The patient demographics were age 63.3 ± 7.45 *years*, sex ratio of 22 males to 18 females, disease duration 8.45 ± 2.87 *years*, and levodopa equivalent daily dosage (LEDD) of 323.78 ± 253.58 *mg*/*day*. For this cohort, the off medication UPDRS-III score was 59.63 ± 20.01, the on medication UPDRS-III score was 23.65 ± 17.76 and the on medication, off stimulation UPDRS-III score was 25.00 ± 13.46. The LEDD percent reduction following surgery was 54% ± 31%. The absolute UPDRS-III post-operative improvement was 35 ± 17.

### 2.2 Data Acquisition

QSMs from mGRE data were reconstructed with MEDI-L_1_ ^19^. Pre-surgical UPDRS-III scores *u* on and off-medication from LCT and post-surgical UPDRS-III scores on-stimulation and off-medication were collected^20^ and the UPDRS-III improvement was computed:

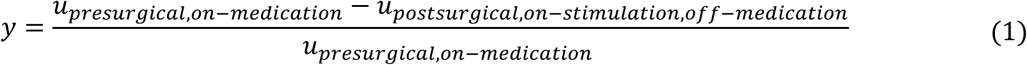

The substantia nigra (SN) and STN were segmented with radiologist supervision. Collected sample size was 40, with 3 cases excluded by motion artifacts. The proposed pipeline is discussed in section *2.3* *Preprocessing* and illustrated in Figure 1.

**Figure 1.**
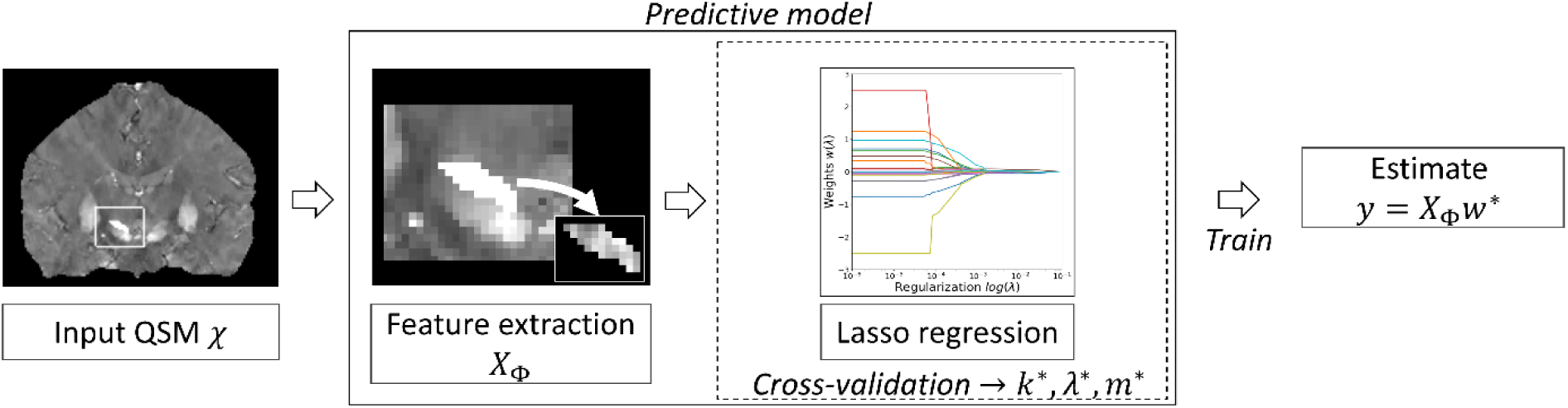
Proposed pipeline with inputs QSM and ROIs. Features were extracted from the input QSM *X* over each ROI (joint entropy feature map shown) and passed to a Lasso regression model after normalization, label noise compensation augmentation, and recursive feature selection. A total of 93 features were extracted over 17 image types available in `PyRadiomics`. Additionally, 14 shape features were extracted on the original image for a total of 1595 features per ROI.

### 2.3 Preprocessing

Figure 1 depicts the proposed pipeline with QSM and ROI inputs. Features were extracted from each ROI (joint entropy feature map shown) and passed to a Lasso model after normalization, label augmentation, and recursive feature selection. A total of 93 features were extracted over 17 image types available in `PyRadiomics`^21^. Additionally, 14 shape features were extracted on the original image for a total of 1595 features per ROI.

### 2.4 Model

A linear regression with Lasso penalization technique implemented with SciKit-Learn was performed to predict UPDRS-III improvement outcome using 1595 features per ROI^22^.

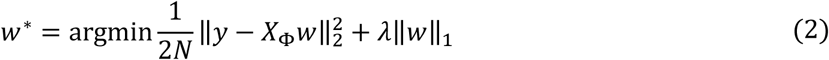

Here, UPDRS-III improvement outcomes *y* are predicted using optimal weights *w*^∗^ and test features *X̂*_Φ_, *λ* is a regularization parameter, and *N* = 37 denotes the number of patients. Given the dataset size, a leave-one-out approach for subject-specific models^23^ was used with each patient withheld and remaining *N* − 1 patients were used for training.

### 2.5 Noise Compensated Data Augmentation

The Lasso training dataset can be augmented using computed label noise statistics. In this work, a Gaussian distribution was fit to the *N* − 1 original UPDRS-III improvements as 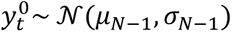, with mean *μ*_*N*−1_ and standard deviation *σ*_*N*−1_. For each model with original features 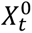, noise instances *∈*_*q*_ were drawn from the Gaussian distribution *N*(0, *zσ*_*N*−1_), with *z* = 1.96, and added to samples in the dataset 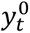:

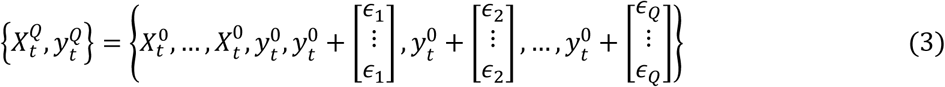

This newly augmented dataset of size (*Q* + 1)(*N* − 1) with labels 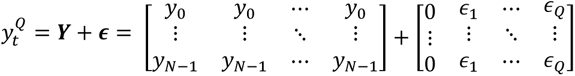. Notice the original labels are preserved in the first column. Here, the use of this augmentation was referred to as “label noise compensation.” For comparison, Lasso was trained with other data augmentation strategies including bootstrap, wild bootstrap^24^, and synthetic minority oversampling with Gaussian noise (SMOGN)^25^.

### 2.6 Training

Nested cross-validation was used to train and test models for each subject. In the outer loop, the model was trained with *N* − 1 subjects and tested with one subject. For the inner loop, *k*-fold cross-validation was performed.

Recursive feature selection was used to reduce the optimal number of normalized features *m*^∗^ from all ROI features *p* = 6380, with step size Δ*m* = 1000 features, for input to Lasso.

Splits *k* = {2,3,4,5}, selected features *m*, and regularization *λ* were chosen by minimizing training error. The minority class (*y*_*train*_ < 0.3) was resampled for stratified cross-validation.

The dataset was augmented (*Q* = 10) with label noise. Least angle regression solver weights (machine epsilon regularization *∈*_*c*_ = 0.1) were initialized at 0. The model was retrained over the entire (*N* − 1) dataset using *m*^∗^ features, split *k*^∗^, and regularization *λ*^∗^ from the inner loop. Training was repeated with standard and noise compensated classifiers.

For each patient, the prediction *ŷ* was a linear combination of features and weights, *ŷ* = *X̂*_Φ_*w*^∗^.

### 2.7 Evaluation

Performance of LCT and Lasso regressors was evaluated using linear regression (Pearson correlation *r*, slope ℳ, intercept *b*, significance *p*) between the observed UPDRS-III improvement and predicted UPDRS-III improvement. Model accuracy was measured by correlation between the observed and predicted outcome. To assess augmentations over different initial noise states, models were evaluated under 10 additional seeds.

Classifiers and regressors were evaluated with receiver operating characteristic (ROC) curves and analogous regression error characteristic (REC) curves, respectively. The frequency of features with nonzero weights were compared for models. The predictive power of a feature is measured by the number of nonzero weights *w*^∗^ across *N* patient-specific models. Feature maps were compared in section *3.2* *Predictive Features* for non-responders (*y* < 0.3) and strong responders (*y* > 0.85), a threshold selected to balance with the number of non-responders.

## 3. Results

### 3.1 Model Performance

Figure 2 illustrates model UPDRS-III improvement predictions. LCT estimates failed to correlate (*r* = 0.12, *p* = 0.48, Figure 2a) with true outcome measured by Equation 1. QSM radiomic predictions correlated with improvements (*r*_*max*_ = 0.75, *p* < 0.001) for noise compensated Lasso, Lasso, bootstrap Lasso, wild bootstrap Lasso, and SMOGN (Figures 2b-2f). The noise compensated Lasso model correctly classified all outcomes: “response” with improvement *y* ≥ 0.3 in the first quadrant or “non-response” in the third quadrant in Figure 1b.

**Figure 2.**
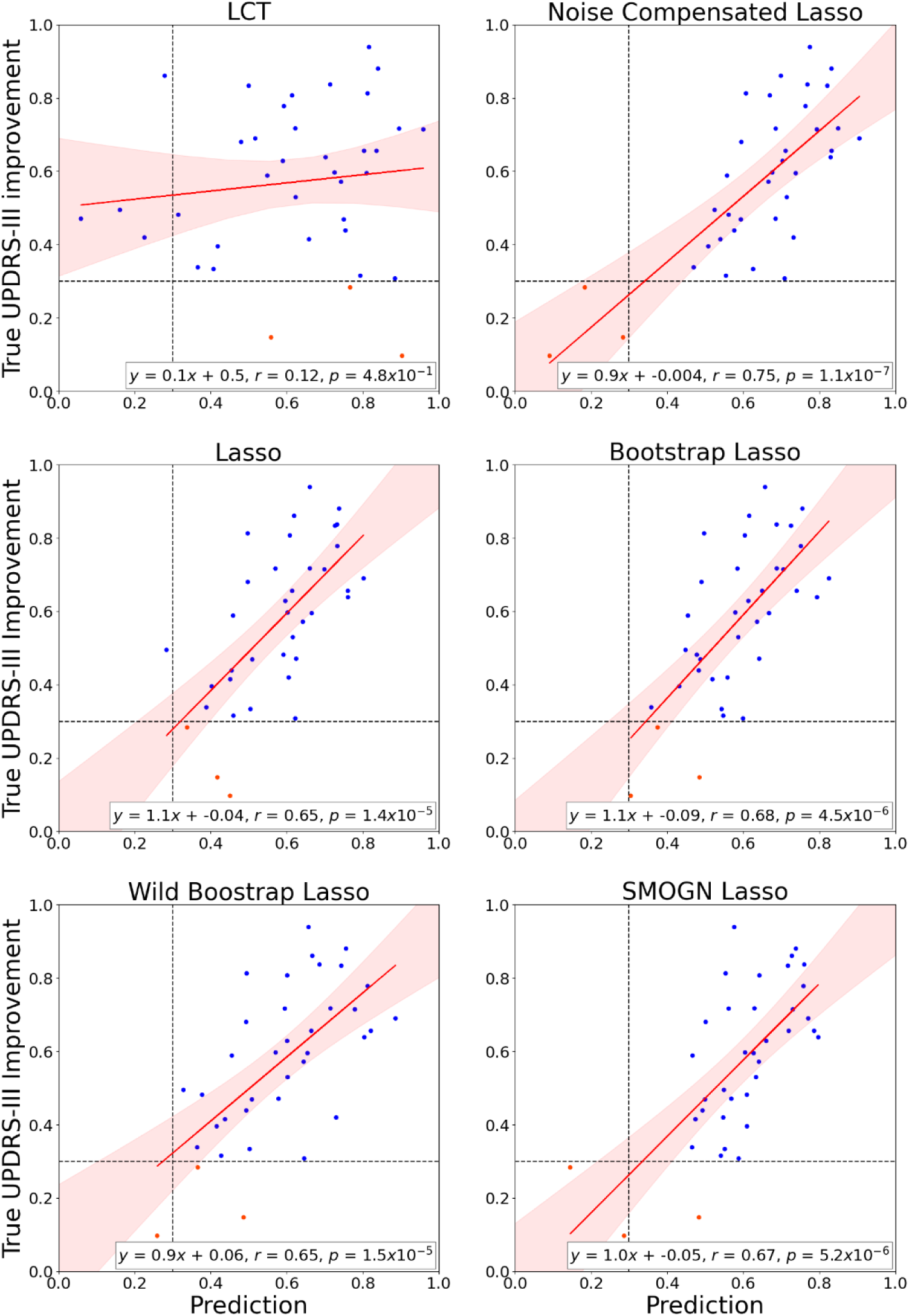
Predicted (horizontal axis) UPDRS-III improvement and true outcomes (vertical axis) using predictors: a) LCT, b) Noise Compensated Lasso (proposed), c) Standard Lasso, d) Bootstrap Lasso, e) Wild Bootstrap Lasso, f) SMOGN Lasso. Blue points indicate a responsive surgical outcome (responder) while red points indicate a non-responsive surgical outcome (non-responder). Note the noise compensated Lasso model correctly classifies all cases.

Noise compensation also increases classifier ROC-AUC from 0.86 to 0.99 (Figure 3a). The regressor REC measures accuracy (vertical axis) of a regression estimator at allowable error bound (horizontal axis) described by the absolute deviation |*y* − *X*_Φ_*w*|. Regressor REC-AUC was 0.82 for radiomics models and 0.73 for LCT (Figure 3b). Note the radiomics models outperform the mean reference estimator, unlike LCT.

**Figure 3.**
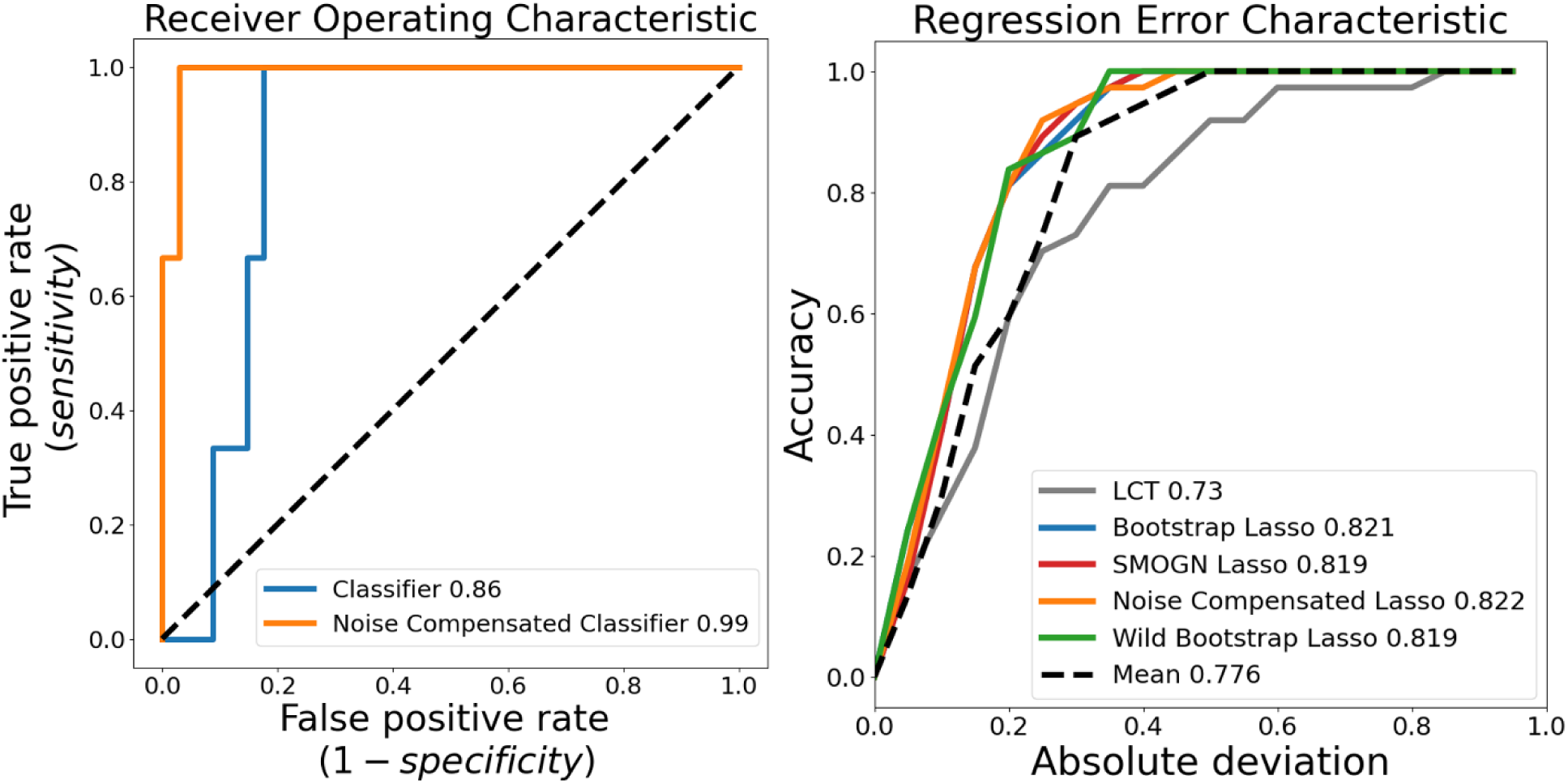
Comparison of classifiers with and without noise compensation (a) and regression estimators with various data augmentation during training (b). The ROC shows an improvement of Δ*AUC* = 0.13. The REC measures accuracy (vertical axis) of a regression estimator at allowable error bound (horizontal axis) described by the absolute deviation |*y* − *X*_Φ_*w*|. Note the radiomics models outperform the mean reference estimator, unlike LCT.

When noise compensated, wild bootstrap, and SMOGN results were fit over the additional noise states, the median result over all fits was found and the regression correlation coefficient approached approximately *r* ≈ 0.70 for each augmentation strategy. The multi-noise state model performances were (*r* = 0.73, ℳ = 1.15, *b* = −0.1, *accuracy* = 0.97 for noise compensated Lasso, (*r* = 0.70, ℳ = 1.12, *b* = −0.07, *accuracy* = 0.97) for SMOGN Lasso, and (*r* = 0.68, ℳ = 0.94, *b* = 0.03, *accuracy* = 0.95) for wild bootstrap Lasso. All correlations were highly significant (*p* < 0.0001).

### 3.2 Predictive features

The most predictive features for the considered ROIs were found on the QSM Coiflet wavelet decomposition of the STN and the gray level dependence matrix (GLDM) from the SN QSM (Figure 4). Specific features were SN large dependence high gray level emphasis (a measure of coarseness) and STN gray level size zone matrix (GLSZM) entropy shown in Figure 5 for strong responders (*y* > 0.85) and non-responders (*y* < 0.3). SN coarseness was higher in non-responders (0.92) versus responders (0.65). Responder STN wavelet entropy was higher (0.61) versus non-responders (0.52). However, neither difference was significant (*⍺* = 0.05).

**Figure 4.**
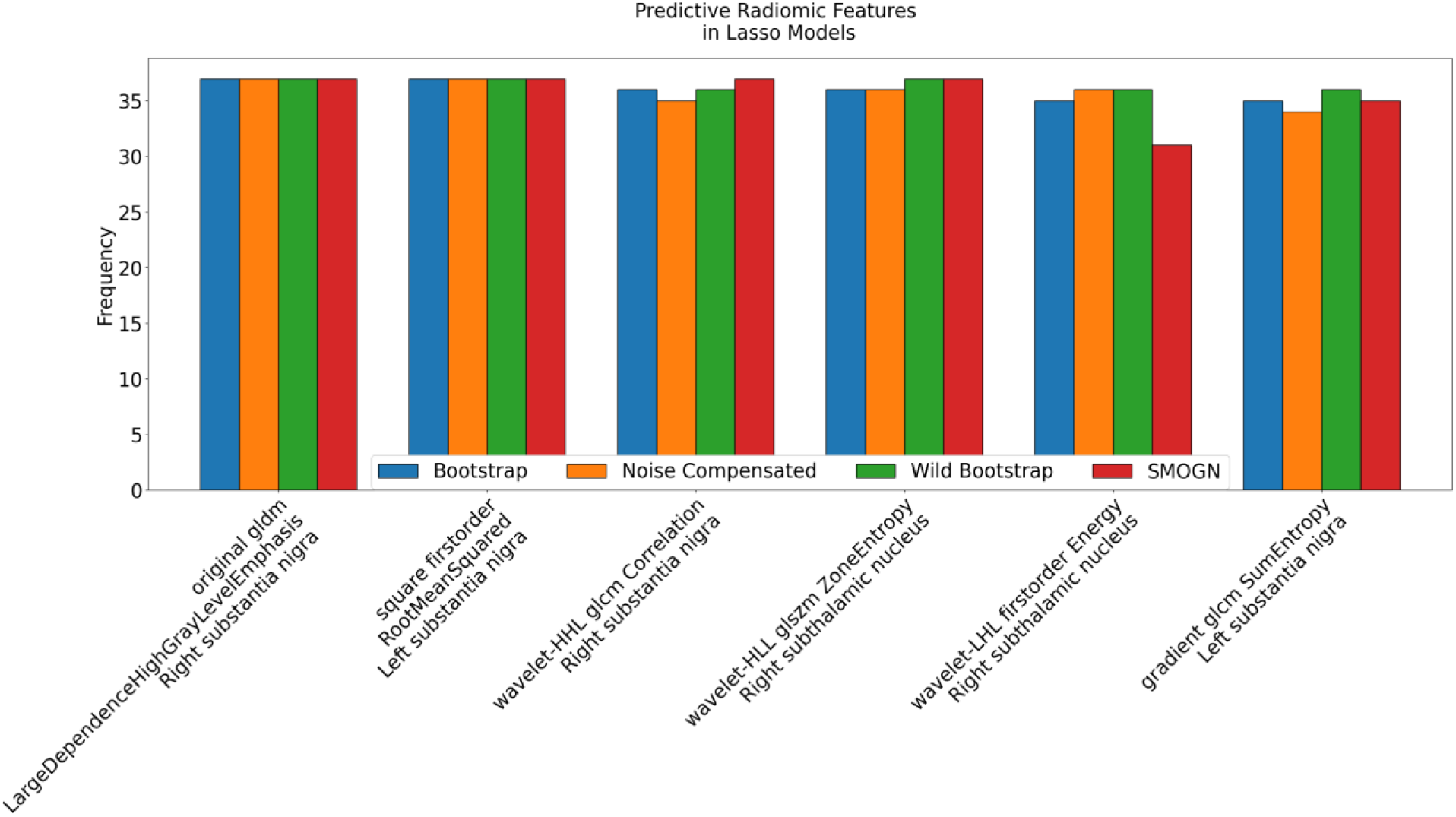
Predictive radiomic features sorted by frequency in *N* = 37 models. All models use the first feature, GLDM large dependence high gray level emphasis, which measures the coarseness of the substantia nigra. In the subthalamic nucleus, the wavelet GLZSM entropy is most predictive, which measures the heterogeneity of the susceptibility distribution.

**Figure 5.**
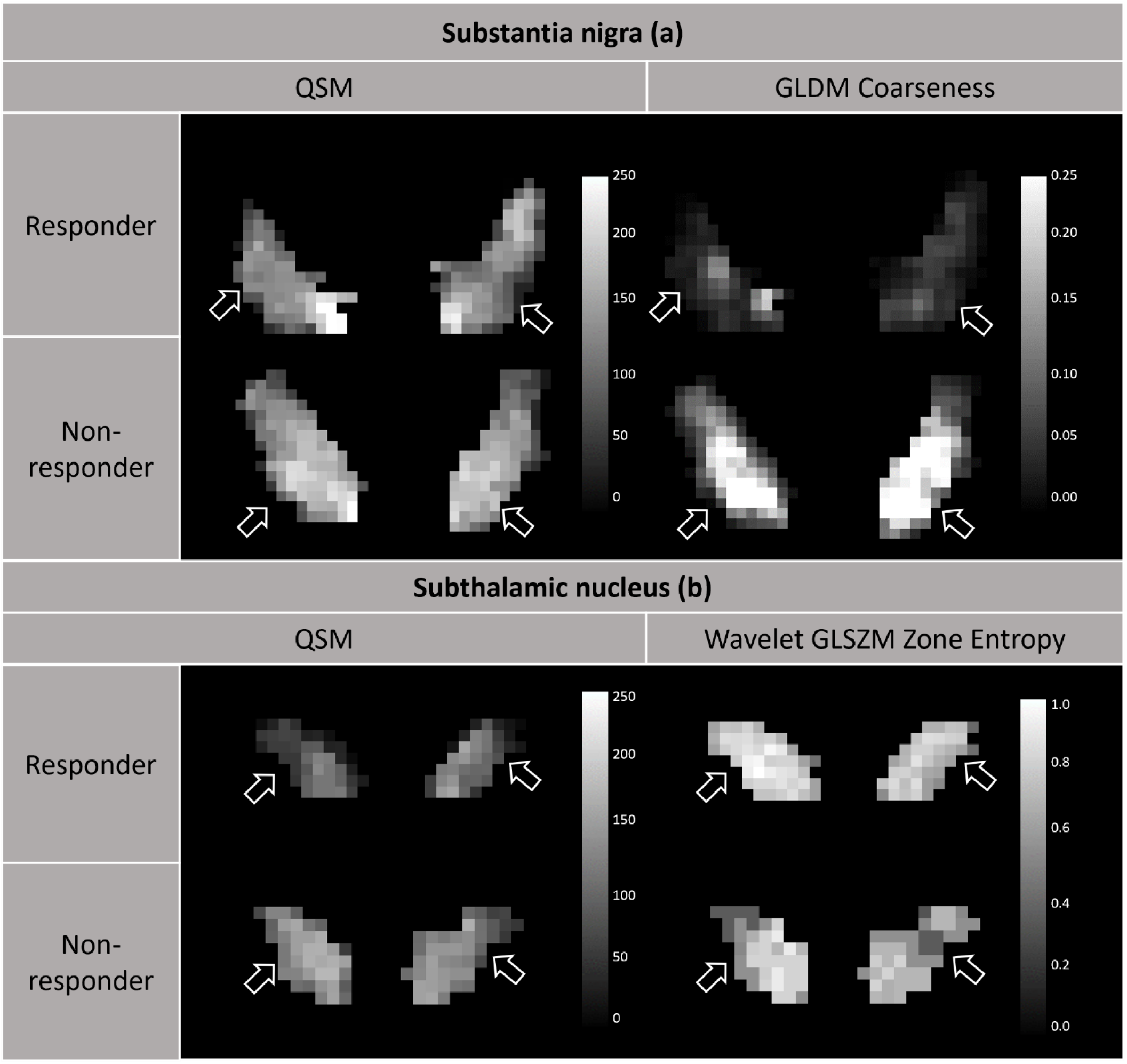
QSM and normalized predictive feature maps or responders (first row) and non-responders (second row) in the substantia nigra (left, a, GLDM large dependence high gray level emphasis, or coarseness) and subthalamic nucleus (right, b, wavelet GLZSM zone entropy). The increased gray level emphasis (a) and the decreased zone entropy (b) in non-responders versus responders were consistent with the ROI feature trends in the *Predictive features* section of the *Results*.

## 4. Discussion

### 4.1 Summary

All models that used QSM radiomics predict DBS UPDRS-III improvements better than LCT. Particularly, label noise compensated models using QSM radiomics performed the highest prediction accuracy. This QSM radiomic prediction is consistent with previous findings of ROI analyses including textual features on PD symptom burden^13,26^. This study is significant in that a numerical prediction of UPDRS-III improvement is offered from routinely acquired QSM used for presurgical targeting.

### 4.2 Prior Works

There are large uncertainties in LCT predictions^20,27^. LCT fails to predict levodopa-resistant symptoms such as tremor improvement^28^ and certain dyskinesias^29^. Additional inaccuracies in LCT result from dose dependence^30^ and disease duration^3^ while inducing burdensome withdrawal symptoms^28^.

Magnetoencephalography^31^ and transcranial direct current stimulation (tDCS)^32^ both demonstrated correlation with DBS improvement, yet neither is routinely acquired in pre-surgical planning. MRI-based brain morphology and functional connectomes have been used to predict DBS motor outcome with limited accuracy^33–37^. QSM from routinely acquired MRI provides predictive features from deep gray nuclei iron distributions. SN texture features reflect degeneration of nigral neurons, reducing dopamine receptor stimulation where iron is a cofactor in dopamine synthesis^38^. Loss of GABAergic inhibition in the STN results in iron deposition encoded by texture features. Dysregulation of SN and STN iron disrupts locomotor activation and inhibition^39^, and subsequent motor symptoms measured by UPDRS-III.

Of the compared augmentations, wild bootstrap is the most similar method to noise compensation. Though it improves non-responder estimates, modeling label variability in noise compensation is more effective. The Lasso model provides a significant and accurate prediction of DBS improvements using routinely acquired QSM under typical levodopa regimen.

### 4.3 Model Interpretation

The prediction is a weighted sum of relevant radiomic features in UPDRS-III improvements. Frequency of nonzero weights across patient-specific models indicate features important in outcome prediction. Comparison of responder and non-responder feature maps demonstrate distinction in both the STN and SN. As iron deposition in the SN is associated with PD progression^40^, predictive feature SN coarseness, or prevalence of high susceptibility regions, suggests elevated iron, advanced tissue damage, or less viable circuitry. This finding is supported by DBS patient selection guidance that considers “very late stages” of PD to be an impediment to symptom improvement^41^. Given the reports of SN iron accumulation and disease progression in PD^42^, the association between the SN coarseness feature and non-responders may support this guidance. Also, loss of QSM STN gradient, captured in predictive wavelet entropy features^21^, correlates with loss of motor function^26^, which is consistent with the wavelet decomposition features identified in previous work^14^. These findings affirm both the observation that inhomogeneity of the STN iron distribution and elevated susceptibility in the SN iron distribution may be predictive of motor symptom severity and patient responsiveness to DBS. Yet, these features alone are insignificant between responders and non-responders, underscoring the need for multivariable models such as Lasso for patient selection.

### 4.4 Limitations

This work has several limitations. As a feasibility study, further validation is required. DBS eligibility requires LCT improvement, generating bias in the dataset against patients without LCT improvement who benefit from DBS and precluding healthy subjects entirely. Additionally, the sample size is small compared to the number of features. Finally, the Gaussian distribution is chosen for label noise modeling but may oversimplify the UPDRS-III interval scale.

## 5. Conclusion

In summary, QSM can provide predictive radiomic features to improve DBS patient selection and better outcome prediction over LCT, in addition to assisting in presurgical targeting. Alongside motor symptoms, non-motor predictions may be further explored (apathy scales, linked to microstructure^43^ and depression inventories). The patient population in this work featured STN-DBS, but the ventral intermediate nucleus and globus pallidus pars interna are also well-visualized on QSM for pre-surgical targeting and should be considered in future studies^44^. Beyond deep gray nuclei, whole brain ROIs may be included^45^. Label noise distributions should be investigated to reflect multiple raters present in the clinical setting.

## 7. Data Availability Statement

Code and data will be available upon publication at: https://github.com/agr78/RadDBS-QSM.

## 8. Author Contributions

**AGR:** Conceptualization; Formal analysis; Software; Validation; Visualization; Original draft. **JZ:** Conceptualization; Formal analysis; Software; Validation; Visualization; Review & editing. **CT:** Formal analysis; Validation; Visualization; Review & editing. **DR:** Conceptualization; Review & editing. **SA:** Data curation; Review & editing. **HK:** Formal analysis; Validation; Visualization, Review & editing. **MRS:** Resources; Supervision; Review & editing. **PS:** Conceptualization; Formal analysis; Supervision; Review & editing. **JL:** Funding acquisition; Resources; Supervision; Review & editing. **YW:** Conceptualization; Funding acquisition; Resources; Supervision; Review & editing. **XW:** Data acquisition; Review & editing. **BHK:** Investigation; Methodology; Resources; Supervision; Writing - review & editing.

